# Choroid Plexus Enlargement is Associated with Disease Severity and Elevated White Matter *Myo*-inositol in Progressive Multiple Sclerosis

**DOI:** 10.64898/2026.06.29.26356824

**Authors:** Sneha Senthil, Flavie E. Detcheverry, Samson Antel, Douglas L. Arnold, Jamie Near, AmanPreet Badhwar, Sridar Narayanan

## Abstract

**Introduction:** Choroid plexus (CP) enlargement on brain MRI has been identified as an emerging neuroinflammatory biomarker in multiple sclerosis (MS), yet its relationship to downstream parenchymal neurochemical abnormalities remains unknown. Proton magnetic resonance spectroscopy (^1^H MRS) enables non-invasive in vivo quantification of neurometabolites, making it well-suited to probe downstream consequences of CP pathology in MS.

**Methods:** Ultra-high-field 7T ^1^H MRS was performed in 45 people with MS (pwMS) (28 Relapsing Remitting MS, RRMS; 17 Progressive MS, PMS) and 43 age- and sex-matched healthy controls (HCs) in the posterior cingulate cortex (PCC) and centrum semiovale white matter (CSWM). CP volume, EDSS, and MS Functional Composite measures were also acquired. Group differences in metabolite concentrations were evaluated using Mann-Whitney U tests with correction for multiple comparisons, and associations between CP volume, altered metabolites, and clinical disability and functional measures were investigated.

**Results:** *Myo*-inositol (mI) was significantly elevated and total *N-*acetylaspartate was reduced in both MS subtypes, in the CSWM. In PMS, CP volume was positively associated with CSWM mI/total creatine (tCr) (ρ = 0.63, *p* = 0.008), an association absent in RRMS. Across the combined MS cohort, CP volume correlated significantly with EDSS (ρ = 0.40, *p* = 0.006).

**Conclusions:** WM mI/tCr was elevated and tNAA/tCr was reduced across MS phenotypes compared with controls, reflecting a dual metabolic signature consistent with concurrent glial overactivation and neuroaxonal compromise. Increased CP volume was associated with greater neurological disability across MS phenotypes. The association of CP enlargement with CSWM mI/tCr in PMS suggests a potential link between CP-mediated periventricular inflammation and progressive WM glial pathology. Collectively, these findings support CP volume as a clinically relevant, non-invasive biomarker and restoring CP integrity as a potential therapeutic target in PMS, where effective treatments remain limited.

**Summary statement:** Choroid plexus enlargement associates with periventricular glial activation specifically in progressive MS, and elevated myo-inositol across brain regions confirms pervasive neuroinflammation detectable by ultra-high-field ^1^H-MRS.

## Introduction

Multiple sclerosis (MS) is the most common chronic inflammatory disease of the central nervous system in young adults and a major cause of neurological disability, affecting over 2.8 million people worldwide as of 2020 (1) Identifying sensitive biomarkers of neuroinflammation is therefore important to understand disease progression and to evaluate the efficacy of disease-modifying therapies (DMTs), particularly in progressive MS where treatment options are limited (2).

Proton magnetic resonance spectroscopy (^1^H-MRS) offers a non-invasive means of quantifying neurometabolites *in vivo*, providing biochemical information that complements structural imaging. Acquisition at higher field strengths (such as 7 Tesla [T]) confers particular advantages, including substantially improved signal-to-noise ratio (SNR) and greater spectral resolution between overlapping resonances. This enables more reliable quantification of metabolites that are difficult to resolve at clinical field strengths.

Several neurometabolites measurable by ^1^H-MRS reflect distinct aspects of MS pathology (3). Specifically, studies have reported an increased *myo*-inositol-to-total-*N*-acetylaspartate ratio (mI/tNAA) in the white matter (WM) of MS participants, particularly around the edges of white matter lesions (WMLs). This ratio reflects the combined contribution of increased mI, a marker of glial activation or gliosis, and reduced tNAA, a marker of neuroaxonal injury or loss (4). mI, primarily synthesised in astrocytes, is considered a marker of glial proliferation and neuroinflammation, with increased concentrations observed in WMLs and, to a lesser extent, in normal-appearing tissue in MS (5). Glutamate (Glu), the principal excitatory neurotransmitter, is elevated in acute MS lesions and normal appearing WM (NAWM), likely reflecting increased glutaminase activity and Glu release by activated microglia and macrophages (6, 7).

Despite insights gained from these ^1^H-MRS derived metabolic markers, several knowledge gaps persist in the MS literature. While elevated mI and reduced tNAA in periventricular and WM regions are well-established findings, the biological mechanisms leading to diffuse neuroaxonal injury and glial proliferation have not been fully characterised. One candidate mechanism that has attracted growing attention is inflammation of the choroid plexus (CP), a structure anatomically positioned at the interface between the peripheral immune system and the central nervous system (CNS), and increasingly recognised as an active orchestrator of neuroinflammation rather than a passive conduit.

The CP is located within the brain’s ventricular system and has a role in mediating neuroinflammation in MS. It is a critical site of immune signalling and is responsible for forming the blood-CSF barrier (BCSFB) and regulating cerebrospinal fluid (CSF) composition. In MS, inflamed CP epithelial cells can recruit peripheral monocytes and macrophages into the CSF, enabling leukocyte infiltration through the CP. This may be a key mechanism driving a surface-in gradient of pathology from the ventricular surface into the brain parenchyma (8). BCSFB integrity is compromised as a result, enabling additional immune cells to penetrate the CNS and induce inflammatory changes in oligodendrocytes (9).

CP enlargement has emerged as a clinically relevant imaging biomarker in MS (10, 11). Meta-analysis across 27 studies including over 1,300 MS participants consistently demonstrate larger CP volume relative to HCs, and this carries both diagnostic and prognostic significance (12). Diagnostically, CP volume significantly differentiates MS from its mimics, including Neuromyelitis Optica Spectrum Disorder (13). Prognostically, CP enlargement at baseline predicts future periventricular neurodegeneration, as reflected by the accumulation of atrophied T2-lesion volume, particularly in PMS (14). Beyond structural damage, higher head-size-corrected CP volume is a notable imaging marker of neuroinflammation that is independent of oligoclonal band status and humoral immunity (15), implicating a role in innate immune-mediated rather than B-cell-driven pathology. Taken together, these findings establish CP enlargement as an upstream, mechanistically grounded marker of CNS inflammation in MS. Understanding whether and how CP enlargement relates to markers of glial activation (mI), neuroaxonal loss (tNAA), oxidative stress (GSH), and excitatory/inhibitory imbalance (Glu) could provide important insight into whether these metabolic signatures are independent of, or partly mediated by CP-related neuroinflammatory processes. The present study, therefore, aimed to identify (a) altered brain metabolites in MS compared to age and sex-matched healthy controls using ultra-high-field (7 Tesla) MR spectroscopy and (b) assess the relationship of CP enlargement with levels of altered metabolites and measures of clinical disability and functional impairment.

## Methods

### Participants

Forty five MS participants (28 relapsing-remitting MS [RRMS, 14F], 17 progressive MS [SPMS, 12F]; EDSS=3 [1-7]; age = 51.5 ± 9.25), followed at the MS Clinic of the Montreal Neurological Institute-Hospital, and 41 age- and sex-matched HCs (age = 51.7 ± 10.9; 23F) were recruited for this study. MS participants met the revised McDonald criteria (11), were ≥ 18 years old, and had been untreated or on stable DMT ≥ 6 months. Exclusion criteria were other neurological disorders, recent relapse or steroid treatment < 30 days, claustrophobia, and MRI contraindications. Healthy controls had no history of neurological, psychiatric, or inflammatory conditions. Study size was determined by participant availability and data quality. All study procedures were approved by the institutional ethics board, and all participants gave written informed consent.

### Functional testing

Functional assessment was performed using components of the MS Functional Composite (MSFC) (16) comprising the Timed 25-Foot Walk Test (T25FWT; ambulation), the Nine-Hole Peg Test (9HPT; upper limb dexterity), and the Symbol Digit Modalities Test (SDMT; cognitive processing speed).

### MR acquisition

All participants were scanned using a 7T whole-body MR scanner (MAGNETOM Terra, Siemens Healthineers, Erlangen, Germany) with an 8 Tx/32 Rx-channel head coil (Nova Medical Inc., Wakefield, MA, USA) in parallel transmission (pTx) mode. The imaging protocol included an MP2RAGE acquisition, with the following sequence parameters: 0.7mm^3^ isotropic resolution, FOV = 240 × 240 × 172 mm^3^, TI1/TI2 = 800 ms/2700 ms, TR/TE = 6000 ms/2.74 ms, acquisition time = 10:14 min. Using these anatomical images as reference, two MRS single-voxels were positioned (Figure 1), one over the posterior cingulate-cortex (PCC), a metabolically active hub region (predominantly grey matter, GM), and the other over the centrum semiovale white matter (CSWM), a WM region prone to MS-related damage. The PCC voxel (30 x 20 x 20 mm^3^) was positioned at midline aligned posterior to the corpus callosum, and the CSWM voxel (30 x 15 x 15 mm^3^) was placed in the left supraventricular WM (Figure 1). MRS spectra were acquired using the STimulated Echo Acquisition Mode (STEAM) sequence with the following parameters: TR/TE = 5000/8 ms; mixing time (TM) = 40 ms; flip angle = 90°; 64 to 96 averages in PCC, and 96 to 128 averages in CSWM; 4096 points; 5000 Hz spectral bandwidth; and acquisition time: 5 to 8 min in PCC, and 8 to 10 min in CSWM. Shimming was performed automatically using the Siemens “brain” B0 shim mode, and VAriable Power and Optimized Relaxations delays method was used for water suppression (17). Non-suppressed water spectra were acquired for eddy current correction and water reference quantification (TR/TE = 5000/8.00 ms; TM = 40 ms; 8 averages). The total MR acquisition lasted about 1 hour 15 minutes.

**Figure 1.**
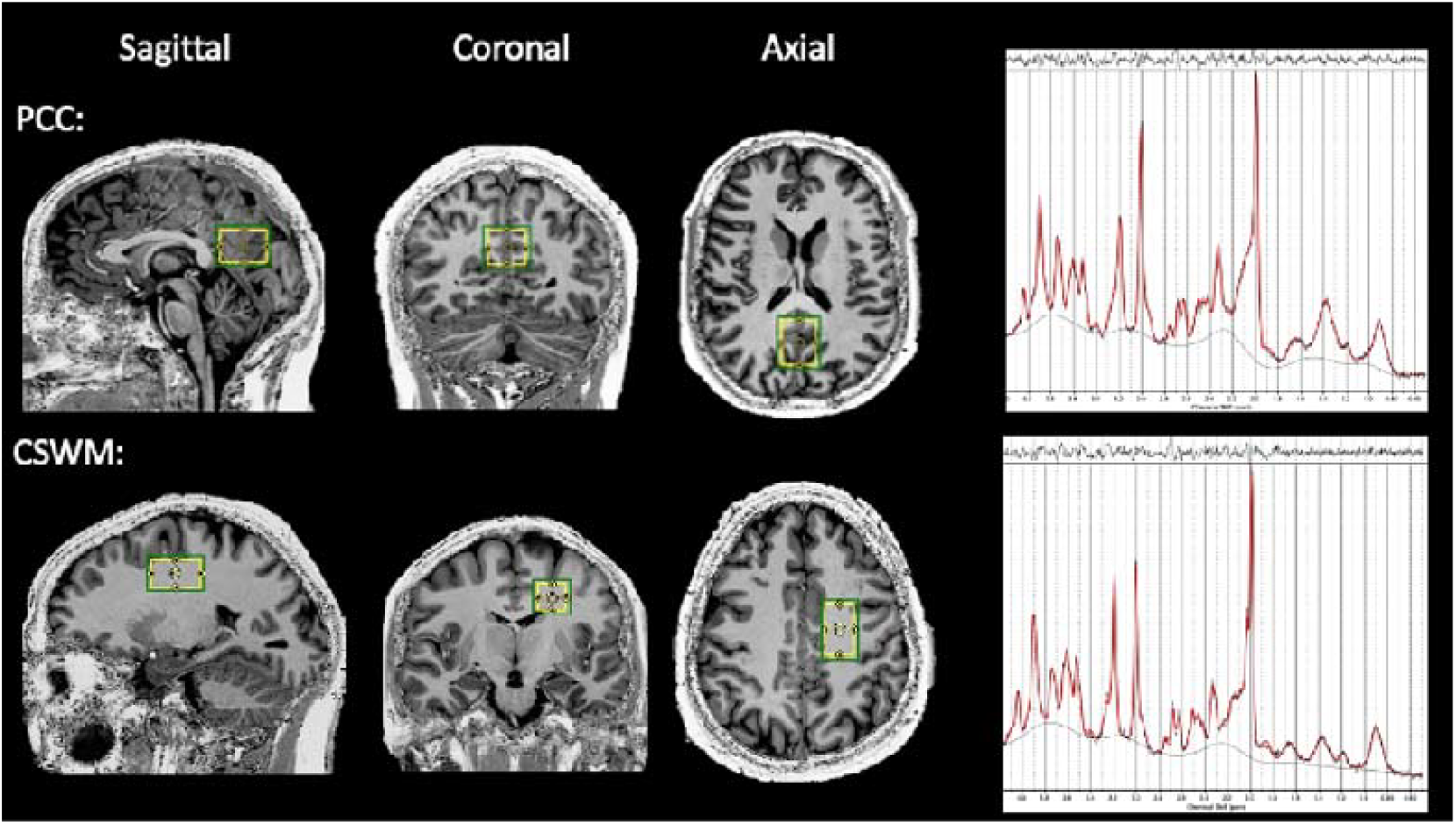
MRS voxels in A) posterior cingulate cortex (PCC) (3 x 2 x 2 (RL x AP x FH) cm^3^); and B) central semiovale white matter (CSWM) (3 x 1.5 x 1.5 (RL x AP x FH) cm^3^) of a representative participant. The green box in the MRI image indicates the shim volume and the yellow box indicates the spectroscopic voxel. MRS spectra from the respective regions are displayed on the right panel.

### MRI processing

T1-weighted images were segmented into GM, WM, and CSF probability maps using Computational Anatomy Toolbox (CAT12) (18). An MRS voxel binary mask was generated in T1 space using the MINC Toolkit (https://bic-mni.github.io), and fractional tissue contributions within the voxel were computed by applying this mask to the tissue probability maps for tissue composition correction.

WMLs in MS participants were segmented from co-registered T1-weighted and FLAIR images using the Lesion Segmentation Toolbox (LST v3.0.0) (19), employing the lesion growth algorithm (κ = 0.3). Total WML volume was extracted per participant and used as a measure of structural lesion burden in subsequent analyses.

### Spectral processing and quantification

The acquired MRS data were preprocessed using the FID-A toolkit (github.com/CIC-methods/FID-A) (20). The preprocessing pipeline performed weighted coil combination (21), alignment of the resulting spectral averages, retrospective frequency and phase drift correction (22), removal of motion-induced bad averages and signal averaging.

Metabolite quantification was performed using the LCModel software (23). The basis set comprised 19 brain metabolites and 9 simulated macromolecule basis spectra. Following expert consensus recommendations (24), quantification of metabolite signal intensity to absolute concentrations was performed (Eq. 1; Supplementary Material S1). This included correction for voxel tissue composition, that is, the relative fractions of GM, WM, and CSF within each spectroscopic volume of interest. T1 and T2 relaxation time constants of water at 7T were obtained from the literature (25, 26). Voxel-specific tissue water concentrations were calculated per participant using Eq. 2 (Supplementary Material S1). Given that our TR >> TE, the T2 relaxation times of metabolites, but not T1, were included from literature (Supplementary Table S1).

Further details of the MRS acquisition and analysis are also included in Supplementary Table S2.

### CP segmentation

CP segmentations were manually performed using MP2RAGE images by a trained rater (S.S., > 3 years of experience). Specifically, the second inversion time (INV2) was used as primary reference for CP identification in line with our previous work (10). Intra-rater variability of ChP segmentation was low, with a coefficient of variation of 4.2% across repeated segmentations performed 3 months apart. Total intracranial volume was extracted using CAT12 applied to the MP2RAGE UNI image, and CP volume was normalised by TIV prior to all statistical analyses.

### Statistical analysis

All statistical analyses were conducted in R (v4.4.1; R Core Team). Major metabolites or metabolite ratios that were included for further analysis included: tNAA (NAA+NAAG), mI, mI/tNAA, tCr (Cr+phospocreatine), tCho (phosphocholine+glycerophosphocholine), GSH, and Glx (glutamate+glutamine). The corresponding ratios to creatine were also included. Normality was tested using the Shapiro-Wilk test, with a significance level of *p* < 0.05 indicating a significant deviation from a normal distribution. Given that the distribution of several metabolites violated the assumption of normality, group differences were evaluated using Mann-Whitney U tests. Pairwise comparisons were performed between HC and RRMS, and HC and PMS groups, separately in the PCC and CSWM regions. To account for multiple comparisons across 13 tests in the RRMS vs HC and PMS vs. HC groups, Bonferroni correction was applied, with statistical significance defined as *p* < 0.00385 (0.05/13).

To examine whether CP enlargement was associated with altered neurometabolite levels, partial Spearman correlations were computed between normalised CP volume (nCPV) and each metabolite that showed a statistically significant group difference, with age and sex included as covariates. Bonferroni correction was applied for the number of metabolites entering the correlation analysis, with the corrected threshold defined accordingly.

CP volume was additionally examined in relation to clinical and imaging measures of disease severity, including WML load, EDSS, and the three MSFC components (T25FWT, 9HPT, and SDMT). These associations were tested using partial Spearman correlations, again adjusting for age and sex, with a Bonferroni-corrected significance threshold of *p* < 0.01 (α = 0.05/5).

## Results

### Characteristics of study population

Table 1 summarizes demographic and clinical characteristics. The MS cohort comprised 28 RRMS (62.2%) and 17 PMS (37.8%) participants. Two RRMS participants had experienced a relapse in the year preceding the study; no recent relapses were recorded in the PMS cohort.

**Table 1.**
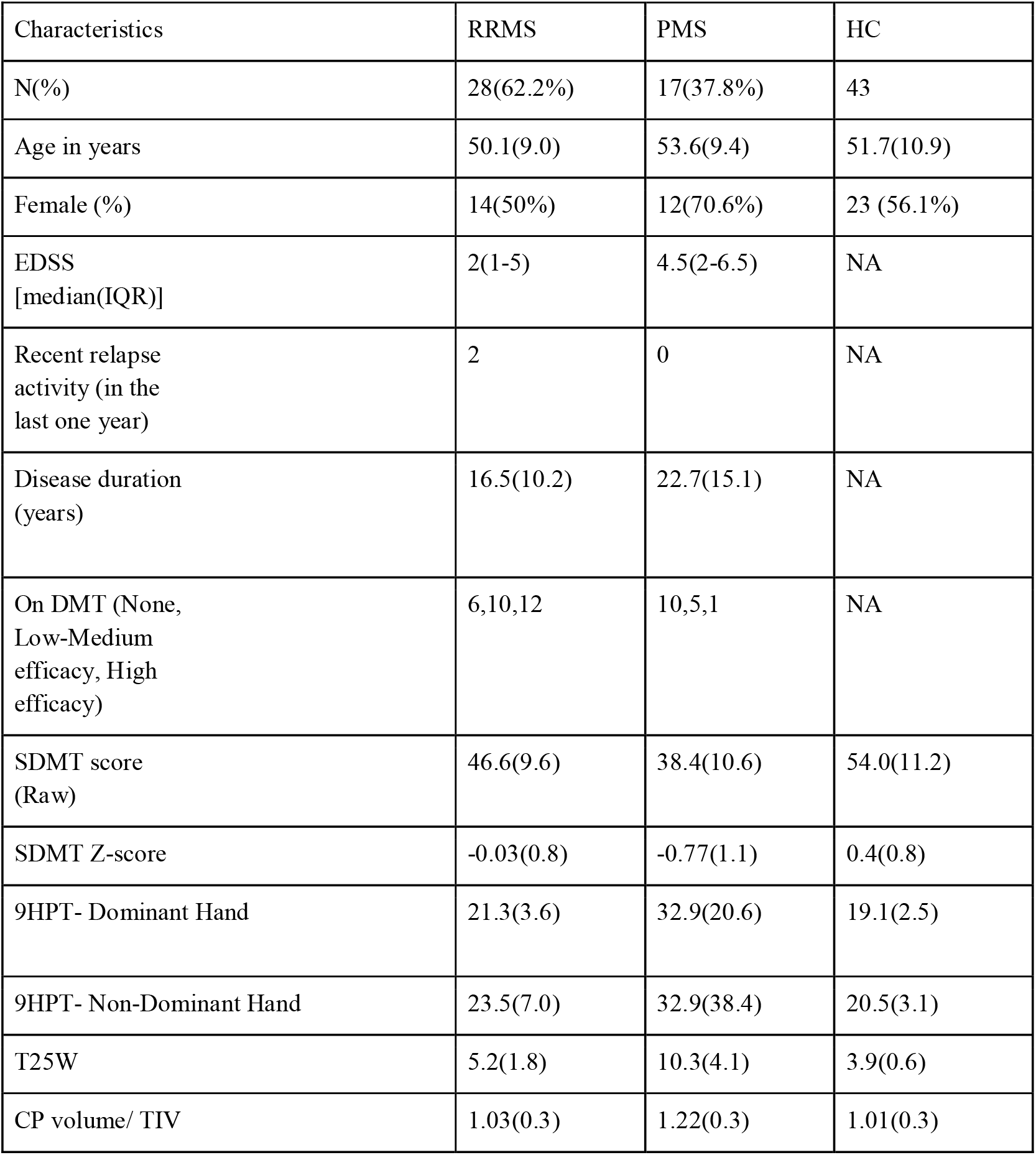
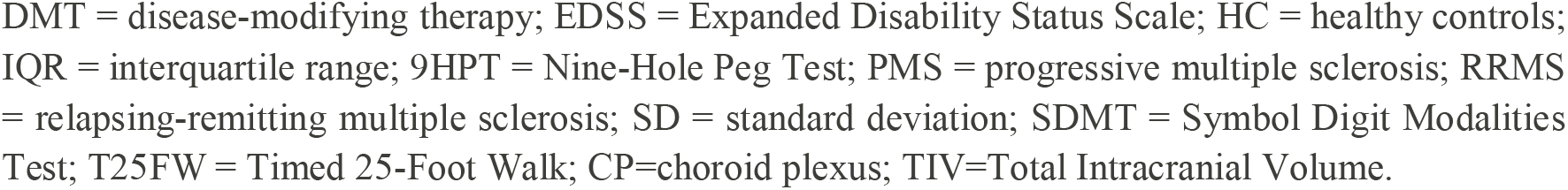
Demographic, clinical, and functional characteristics of the study cohort. All measures are in the format [Mean(SD)] unless indicated otherwise.

| Characteristics | RRMS | PMS | HC |
| --- | --- | --- | --- |
| N(%) | 28(62.2%) | 17(37.8%) | 43 |
| Age in years | 50.1(9.0) | 53.6(9.4) | 51.7(10.9) |
| Female (%) | 14(50%) | 12(70.6%) | 23 (56.1%) |
| EDSS<br>[median(IQR)] | 2(1-5) | 4.5(2-6.5) | NA |
| Recent relapse<br>activity (in the<br>last one year) | 2 | 0 | NA |
| Disease duration<br>(years) | 16.5(10.2) | 22.7(15.1) | NA |
| On DMT (None,<br>Low-Medium<br>efficacy, High<br>efficacy) | 6,10,12 | 10,5,1 | NA |
| SDMT score<br>(Raw) | 46.6(9.6) | 38.4(10.6) | 54.0(11.2) |
| SDMT Z-score | -0.03(0.8) | -0.77(1.1) | 0.4(0.8) |
| 9HPT- Dominant Hand | 21.3(3.6) | 32.9(20.6) | 19.1(2.5) |
| 9HPT- Non-Dominant Hand | 23.5(7.0) | 32.9(38.4) | 20.5(3.1) |
| T25W | 5.2(1.8) | 10.3(4.1) | 3.9(0.6) |
| CP volume/ TIV | 1.03(0.3) | 1.22(0.3) | 1.01(0.3) |

DMT = disease-modifying therapy; EDSS = Expanded Disability Status Scale; HC = healthy controls; IQR = interquartile range; 9HPT = Nine-Hole Peg Test; PMS = progressive multiple sclerosis; RRMS = relapsing-remitting multiple sclerosis; SD = standard deviation; SDMT = Symbol Digit Modalities Test; T25FW = Timed 25-Foot Walk; CP=choroid plexus; TIV=Total Intracranial Volume.

### Group-wise metabolite comparisons

The spectral quality was good across all scans (mean SNR = 25 ± 3.5, FWHM =7.5± 2.4Hz), representative spectra from both brain regions are shown in Figure 1). The major metabolites measured were: mI, tCr, tCho, GSH, Glx, tNAA. For GSH quantification, eight PCC datasets and one CSWM dataset were excluded because of poor spectral fitting quality. Figure 2 presents boxplots of absolute metabolite concentrations and Figure 3 presents tCr-referenced ratios for HC vs RRMS vs PMS. The HC cohorts differed between analyses to match the age and sex characteristics of the RRMS and PMS groups, respectively. Full descriptive statistics are provided in Supplementary Table S3.

**Figure 2.**
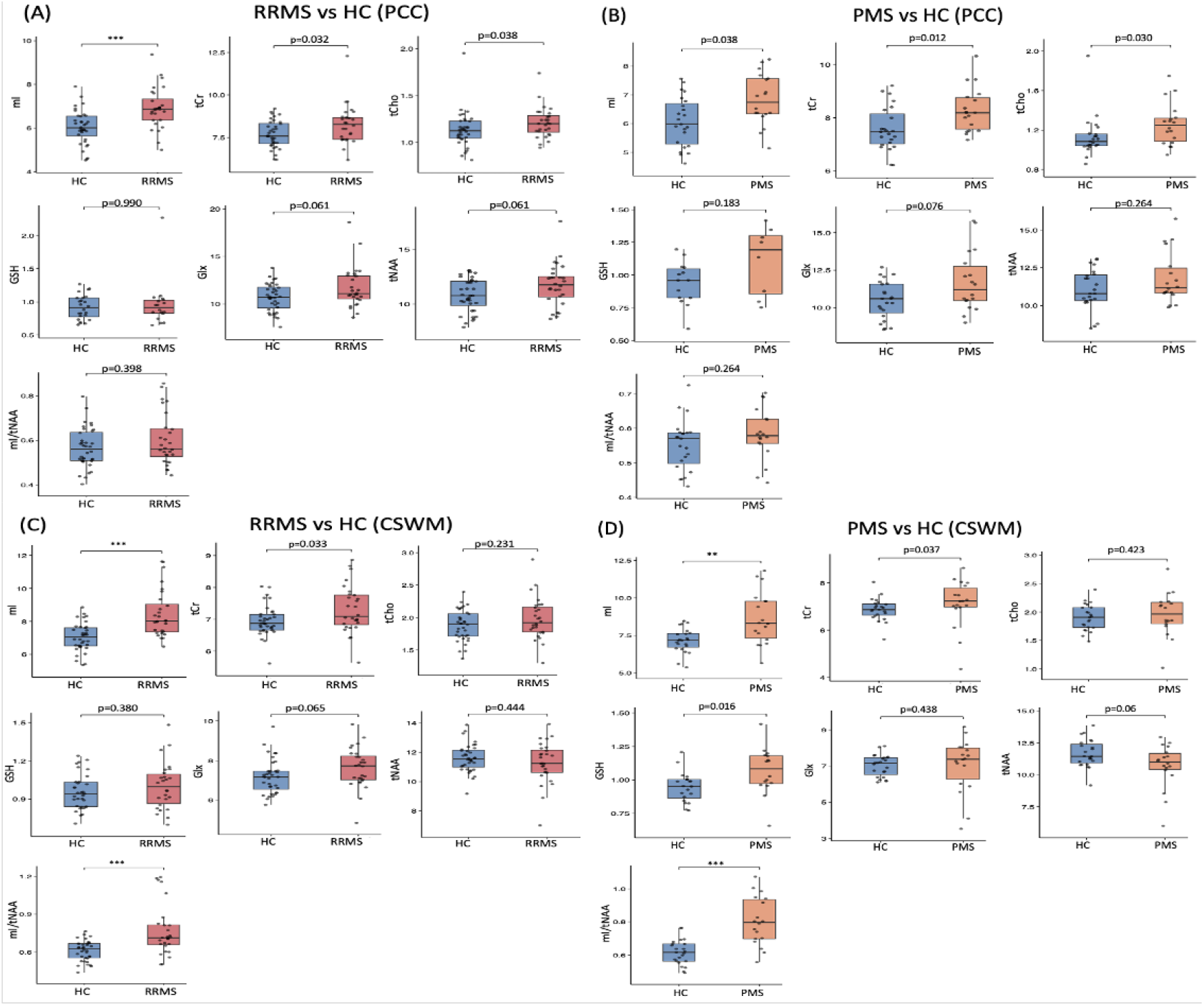
Group comparisons of absolute metabolite concentrations in the PCC and CSWM. Boxplots show (A) RRMS vs HC in the PCC, (B) PMS vs HC in the PCC, (C) RRMS vs HC in the CSWM and (D) PMS vs HC in the CSWM. The asterisk (*) indicates significance in line with the Bonferroni-corrected significance threshold (p < 0.0038).

**Figure 3.**
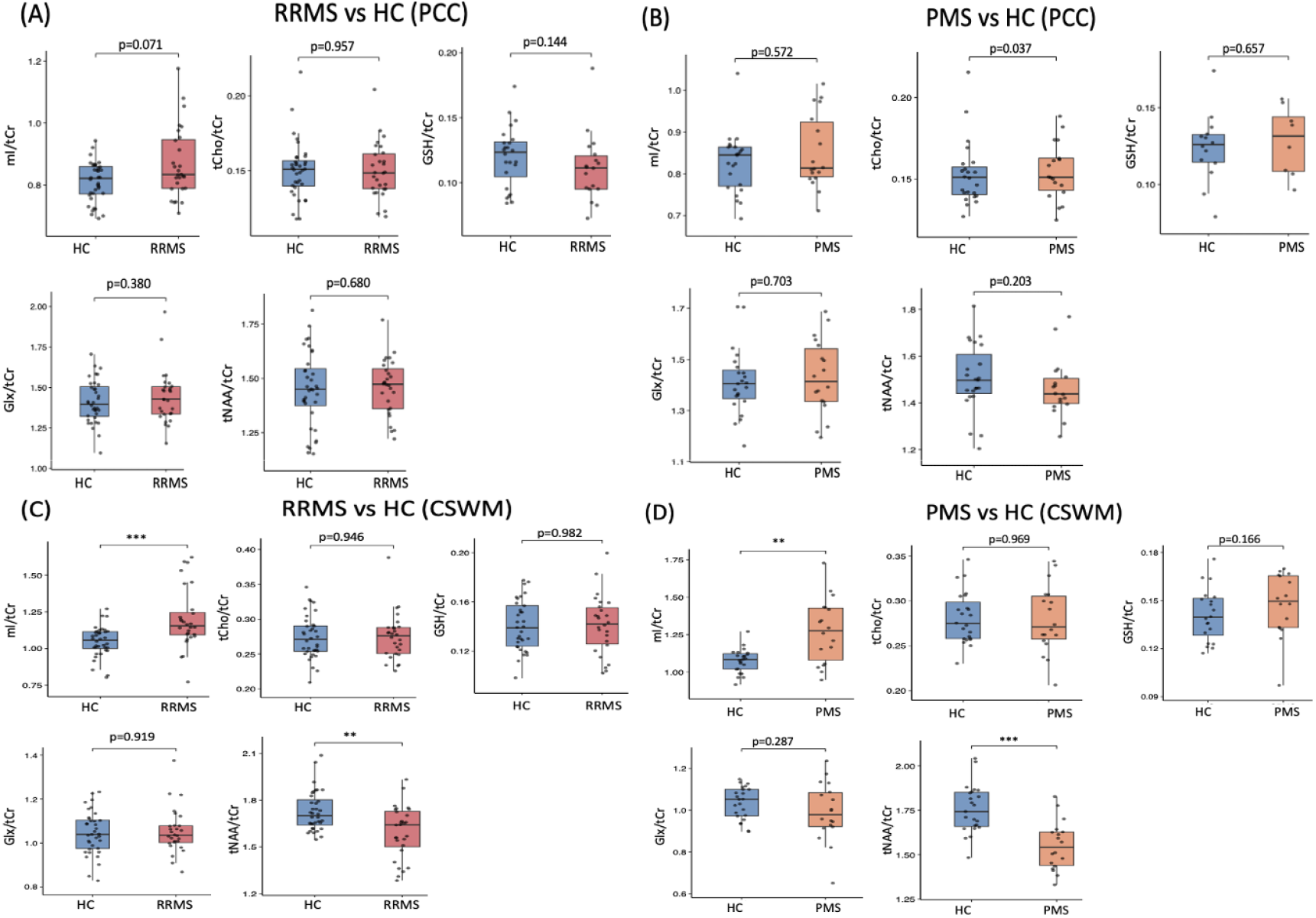
Group comparisons of tCr-referenced ratios in the PCC and CSWM. Boxplots show (A) RRMS vs HC in the PCC, (B) PMS vs HC in the PCC, (C) RRMS vs HC in the CSWM and (D) PMS vs HC in the CSWM. The asterisk (*) indicates significance in line with the Bonferroni-corrected significance threshold (p < 0.0038).

In the PCC, absolute mI concentration was significantly elevated in RRMS relative to matched HC (p < 0.001), whereas no tCr-referenced ratio reached significance. In PMS, absolute mI concentration was nominally higher than HC but did not survive Bonferroni correction, and no significant differences were observed for any tCr-referenced ratio in this region.

In the CSWM, both groups showed significantly elevated mI/tNAA and absolute mI concentration relative to their respective HC cohorts, surviving Bonferroni correction. Among tCr-referenced ratios, mI/tCr was significantly higher and tNAA/tCr significantly lower in both RRMS and PMS compared to HC.

### Association of CP volume with altered metabolite levels

Prior to examining associations with neurometabolite alterations, group differences in nCPV were assessed. nCPV was significantly higher in PMS compared to both RRMS and HCs (*p* < 0.05), and higher in the combined MS cohort relative to HCs (*p* < 0.05). To examine whether CP enlargement was associated with the neurometabolite alterations identified in the group comparisons, partial Spearman correlations were computed between nCPV and metabolite ratios with age and sex included as covariates. In the RRMS group, the altered metabolites were mI in the PCC and mI, mI/tCr and tNAA/tCr in the CSWM. In the PMS group, mI, mI/tNAA, mI/tCr, and tNAA/tCr were altered in the CSWM. Association analyses were restricted to ratios (tCr-referenced ratios and mI/tNAA) since ratio measures are internally normalised and therefore more robust to between-subject variability in voxel placement and receiver gain than absolute concentrations in institutional units (27). Because the RRMS and PMS correlation analyses were conducted independently, Bonferroni correction was applied separately within each group. Accordingly, statistical significance was set at *p* < 0.025 for RRMS analyses and *p* < 0.0167 for PMS analyses. Figure 4 shows the partial Spearman correlation plots for each group. In the RRMS group, no significant associations were found between nCPV and mI in either region (both *p* > 0.025). In the PMS group, nCPV was significantly and positively associated with CSWM mI/tCr (ρ = 0.63, *p* = 0.008). No significant associations were observed for mI/tNAA or tNAA/tCr in this group (both *p* > 0.0167).

**Figure 4.**
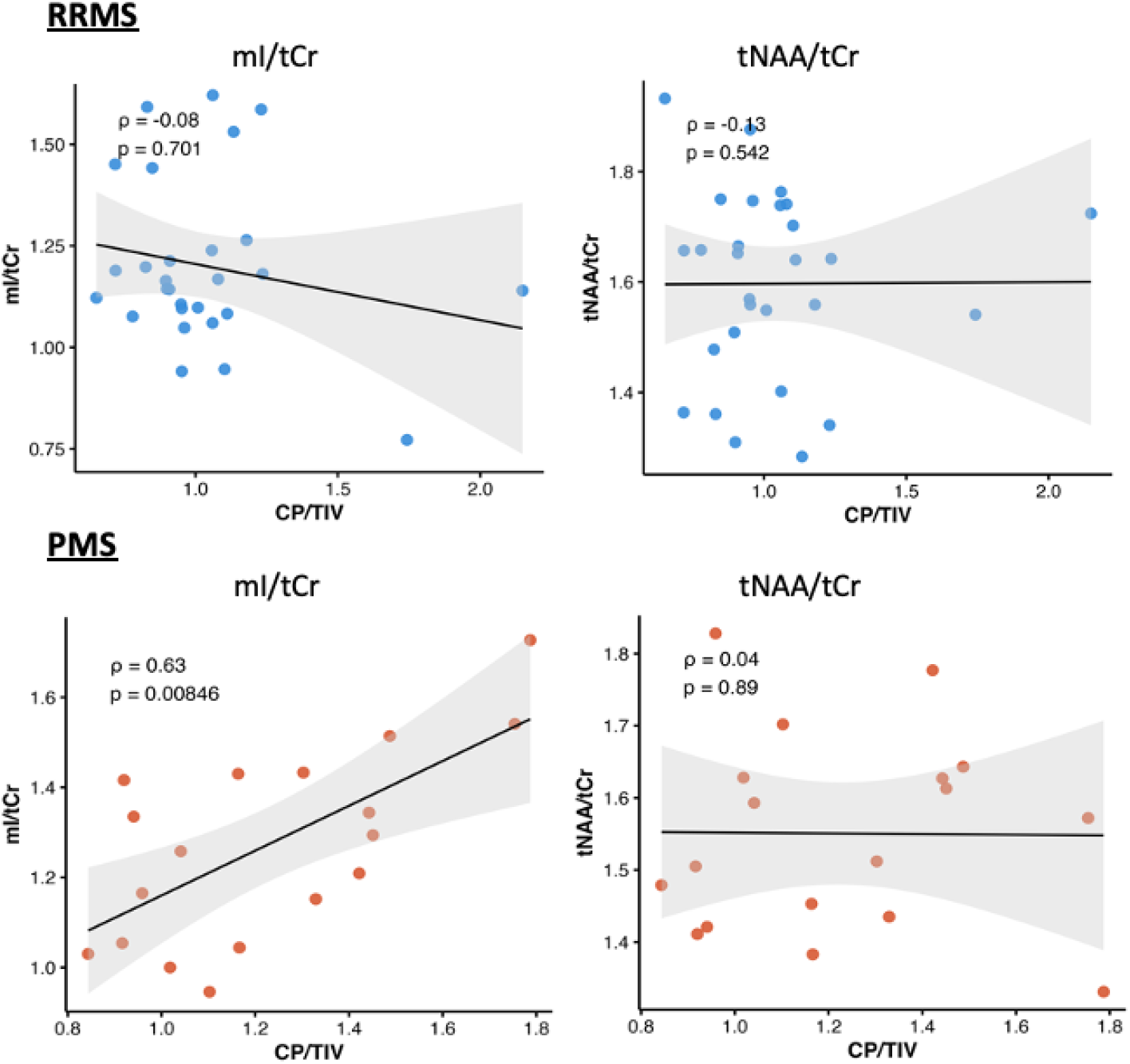
Partial Spearman correlation plots for each group investigating the relationship between CP/TIV volume (x10^3^) vs altered metabolites in the CSWM (mI/tCr, tNAA/tCr), covarying for age and sex. Statistical significance was set at *p* < 0.025 in each group.

### Group differences in functional measures

Figure 5 shows boxplots of MSFC scores for HC vs RRMS vs PMS. Clinical measures were significantly impaired (*p* < 0.0001) in the MS group, indicating deficits in manual dexterity, cognitive processing speed, and mobility. Descriptive statistics are included in Table 1 for each group.

**Figure 5.**
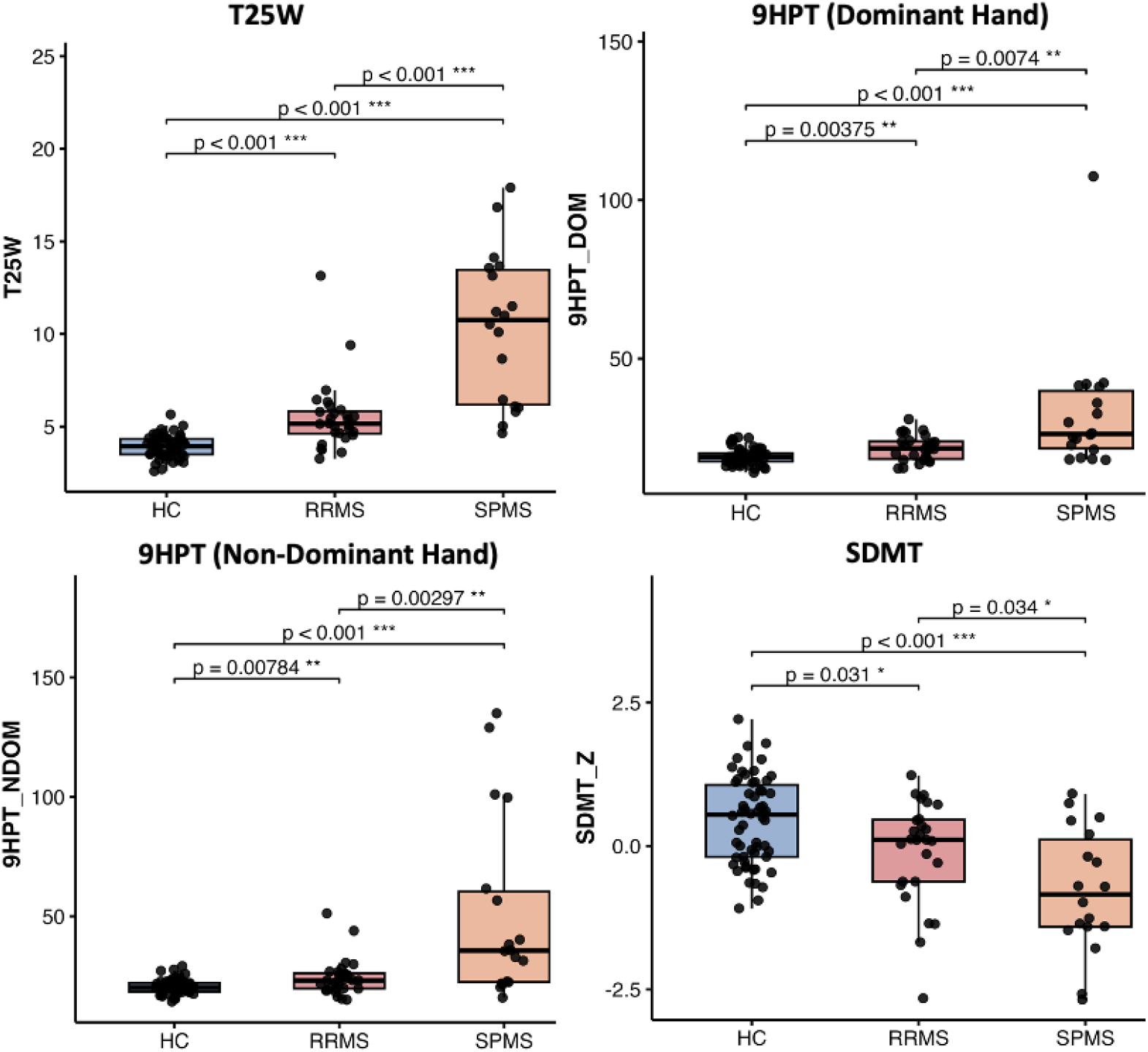
Group differences in MSFC measures among HC, RRMS, and PMS groups. Boxplots illustrate MSFC performance across HC, RRMS, and PMS groups. MS participants demonstrated significantly impaired functional performance compared with HC (*p* < 0.05).

### Association of CP volume with disability metrics

Figure 6 presents partial Spearman correlations between nCPV and clinical and imaging measures in the combined MS cohort, adjusting for age and sex. The following metrics were examined: EDSS, T25FWT, 9HPT (dominant and non-dominant hand), SDMT, and WML volume, with a Bonferroni-corrected significance threshold of *p* < 0.0083 (0.05/6). Across the MS cohort, nCPV was significantly and positively correlated with EDSS (ρ = 0.40, *p* = 0.006), surviving correction for multiple comparisons. A weak positive trend was observed between nCPV and T25FWT (*p* = 0.049); however, this did not survive Bonferroni correction. No significant associations were observed between nCPV and 9HPT (dominant or non-dominant hand), SDMT Z-scores, or WML volume. When RRMS and PMS cohorts were analysed separately, no significant associations were observed between nCPV and any disability metric.

**Figure 6.**
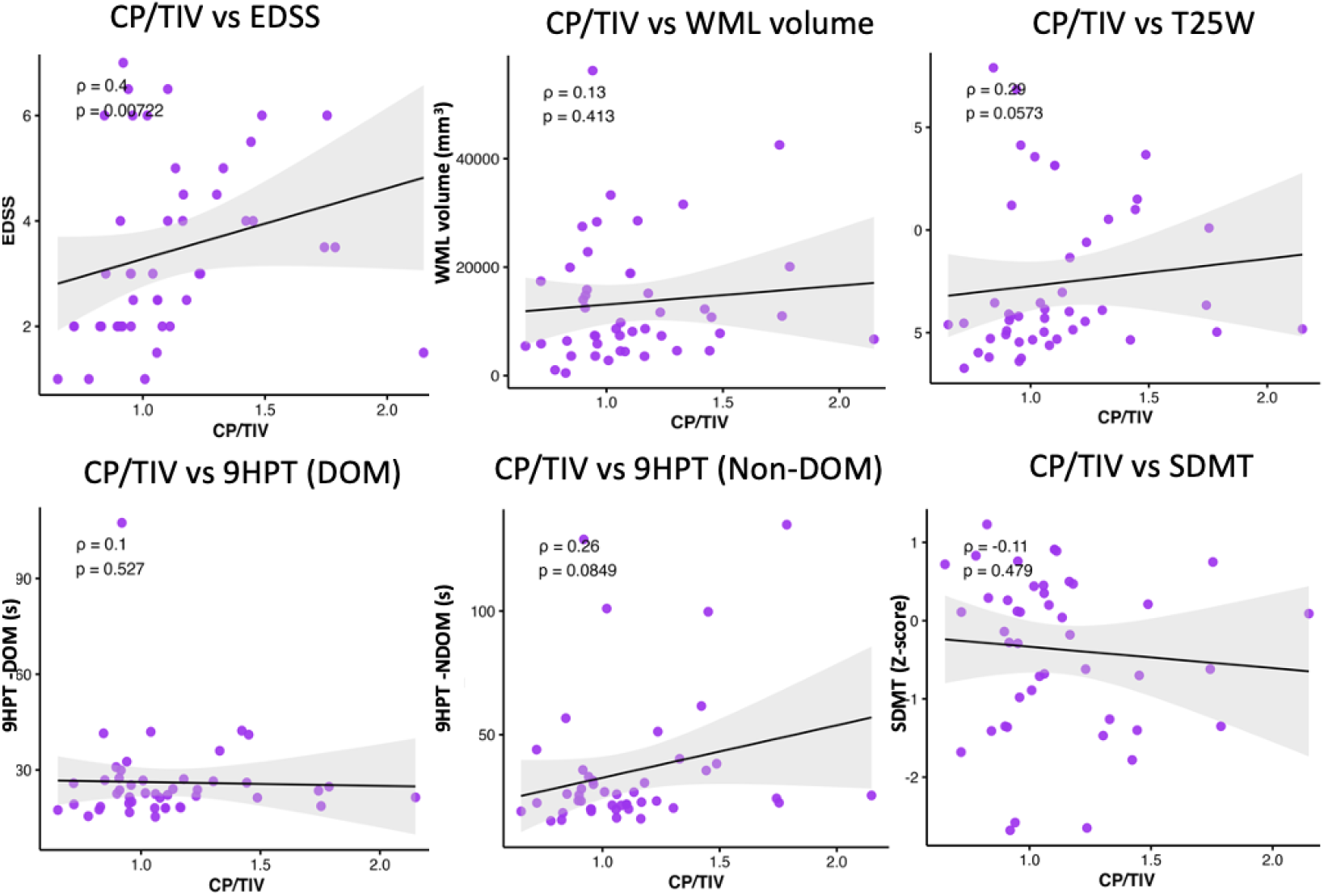
Partial Spearman correlation analyses were performed between CP/TIV (x10^3^) and clinical and imaging metrics, adjusting for age and sex, in the MS cohort. Examined measures included EDSS, WML volume, T25FWT, 9HPT (dominant and non-dominant hand) and SDMT Z-scores.

## Discussion

### Metabolic alterations and underlying pathology

The most consistent metabolic alteration observed across both MS subtypes was elevated mI, a metabolite synthesised primarily in astrocytes that serves as an osmolyte and second messenger precursor (28). As early as the clinically isolated syndrome stage, mI is elevated in NAWM of the centrum semiovale compared to HCs, demonstrating that reactive astrogliosis precedes the accumulation of T2-visible lesions and is an early feature of the disease (29). Quantitative MRS of NAWM in clinically definite MS has consistently shown increased mI concentration alongside increased tCr and decreased tNAA (4). More recently, a longitudinal 7T MRSI study of 20 RRMS patients followed annually for up to three years demonstrated that mI and tNAA in NAWM are sensitive biomarkers of smouldering-associated worsening, with mI changes associating with paramagnetic rim lesions, brain atrophy, and T2 lesion load (30). These *in vivo* MRS findings are directly supported by histopathological evidence linking mI/tCr elevation to histologically confirmed astrogliosis (31).

### CP enlargement and associations with metabolic alterations and disease severity

To our knowledge, this is the first study to examine the relationship between CPV and ^1^H-MRS metabolite alterations in MS. The significant positive association between nCPV and CSWM mI/tCr in PMS is a novel finding with potential clinical implications. The association of mI/tCr with CPV in the PMS subgroup suggests that individuals with greater CP enlargement, reflecting more pronounced CP-mediated neuroinflammation, also exhibit higher levels of periventricular glial activation. If replicated longitudinally, this relationship could inform patient stratification for trials targeting the CP neuroinflammatory pathway. It may also serve as a treatment response biomarker for emerging BCSFB-directed therapies in PMS, where treatment options remain limited.

The association between nCPV and EDSS observed in our combined MS cohort is consistent with and supported by the existing literature (32). A recent systematic review and meta-analysis pooling data from 21 case-control studies in over 1,400 pwMS demonstrated a significant positive association between nCPV and EDSS, confirming CPV as a correlate of multidimensional neurological disability (12). Our somewhat larger effect size (ρ = 0.40) may reflect covariate adjustment for age and sex, improving specificity, as well as the methodological consistency inherent to a single-centre design with standardised CP segmentation.

### Limitations

Several limitations should be acknowledged. The MRS protocol employed single-voxel acquisitions in the PCC and CSWM, rather than whole-brain MRSI coverage. Consequently, metabolite alterations in WMLs and other regions could not be captured. Future studies incorporating whole-brain MRSI are warranted. Absolute concentrations of metabolites relied on assumed relaxation parameters and stimulated MM spectra, both of which may vary with age and disease (33, 34). The cross-sectional design precludes causal inference, and the modest, clinically heterogeneous subgroup sizes warrant cautious interpretation. Finally, secondary PMS and primary PMS participants were combined into a single group, although these subtypes may differ biologically.

## Conclusion

This study demonstrates that ultra-high-field 7T ^1^H-MRS detects significant neurometabolic abnormalities in pwMS, particularly elevated mI within the CSWM. This is indicative of ongoing glial activation and neuroinflammation. CP enlargement was significantly associated with CSWM mI/tCr in PMS, suggesting a potential link between CP-mediated periventricular inflammation and progressive WM glial pathology. The positive association between CP enlargement and disability severity, further supports the clinical relevance of CP pathology in MS. Together, these findings suggest that CP imaging and brain metabolic profiling may provide useful biomarkers for future therapeutic stratification especially in PMS.

## Supporting information

Supplemental File for Review

## Data Availability

All data produced in the present study are available upon reasonable request to the authors.

## Acknowledgements

This work was supported by a Canadian Institutes of Health Research (CIHR) grant (S.N., #153005), the Réseau de Bio-Imagerie du Québec (RBIQ) Pilot Grant, the Centre de Recherche de l’Institut Universitaire de Gériatrie de Montréal (CRIUGM) programme d’appui à des Projets de Recherche Stratégiques et Structurants (PRSS), and the Fonds de Recherche Québec-Santé (FRQS) (A.B); FRQS Chercheurs boursiers Junior 1 and 2, and the Fonds de soutien à la recherche pour les neurosciences du vieillissement from the Fondation Courtois (A.B.); and the FRQS bourse de formation au doctorat (2024) (#352199, F.E.D.; #332672, S.S). We also thank our MRI technologists Ronaldo Lopez, David Costa, and Soheil Mollamohseni Quchani; as well as Rozie Arnautelis for assistance with recruitment of participants.

