## Supplemental File for Review for "Choroid Plexus Enlargement is Associated with Disease Severity and Elevated White Matter *Myo*-inositol in Progressive Multiple Sclerosis"

**Supplementary material S1**: Equations for absolute metabolite quantification

**Equation 1.**

$$C_{abs\_met} = \frac{LCMoutput_{met}}{conc_{H2O} \cdot ATT_{H2O}} \cdot \frac{\left(H2O_{WM} \cdot e^{-\frac{TE}{T2_{WM}}} \cdot \left(1 - e^{-\frac{TR}{T2_{WM}}}\right) \cdot f_{WM} + H2O_{GM} \cdot e^{-\frac{TE}{T2_{GM}}} \cdot \left(1 - e^{-\frac{TR}{T1_{GM}}}\right) \cdot f_{CSF} + H2O_{CSF} \cdot e^{-\frac{TE}{T2_{CSF}}} \cdot \left(1 - e^{-\frac{TR}{T1_{CSF}}}\right) \cdot f_{CSF}\right)}{e^{-\frac{TE}{T2_{met}}} \cdot (f_{WM} + f_{GM})}$$

*With:*

$C_{abs\_met}$: Absolute concentration for metabolite of interest

$LCMoutput_{met}$: LCmodel output for metabolite of interest

$ATT_{H20}$ = 1 (Attenuation factor provided by LCmodel [unitless])

$H20_x$: Assumed visible water concentration for x tissue of interest (WM, GM, CSF) (mmol/L)

$T2_x$: T2 relaxation time for x tissue of interest (WM, GM, CSF) (ms)

TE= 8ms

TR = 5000ms

$f_x$ = Volume fraction for x tissue of interest (WM, GM, CSF) within MRS voxel

concH20= voxel-specific tissue water concentrations, calculated using Equation 2.

**Equation 2:**

$$conc_{H2O} = f_{WM} \cdot [H2O]_{WM} + f_{GM} \cdot [H2O]_{GM} + f_{CSF} \cdot [H2O]_{CSF}$$

Where:

$[H20]_{WM}$ = 36100 mmol/L

$[H20]_{GM}$ = 43300 mmol/L

$[H20]_{CSF}$ =53800 mmol/L

Values of $H20_x$ were derived from Dhamala E, Abdelkefi I, Nguyen M, Hennessy TJ, Nadeau H, Near J. Validation of in vivo MRS measures of metabolite concentrations in the human brain. *NMR Biomed*. 2019;32(3):e4058. doi:10.1002/nbm.4058

**Supplementary Table S2**: **MRS in MRS checklist**

1. **Hardware**

| | |
|---|---|
| Field strength (T) | 7 T |
| Manufacturer | Siemens |
| Model (software version) | Terra (VE12U) |
| RF coils: nuclei (transmit/receive), number of channels, type, body part | 8 Tx/32 Rx head coil |
| Additional hardware | N/A |

2. **Acquisition**

| | |
|---|---|
| Pulse sequence | STEAM |
| Volume of interest (VOI) location | **Posterior cingulate cortex (PCC)**: oblique axial voxel parallel to the line connecting the inferior aspect of the genu and splenium of the corpus callosum (callosal line). Voxel was positioned on the mid-line grey matter posterior to the splenium of the corpus callosum to intersect PCC and precuneus<br>**Central Semiovale white matter (CSWM)**: superior to the lateral ventricles and corpus callosum, subjacent to the cerebral cortex |
| Nominal voxel size | PCC: 30mm x 20mm x 20 mm<br>CSWM: 30mm x 15mm x 15mm |
| TR/TE [ms] | 5000/8ms; TM=40ms |
| Total number of excitations or acquisitions per spectrum | Water suppressed (PCC): 64 to 96 averages<br>Water suppressed (FWM): 96 to 128 averages<br>Non-water-suppressed: 16 averages |
| In time series for kinetic studies<br>i. Number of averaged spectra (NA) per time point<br>ii. Averaging method<br>iii. Total number of spectra (acquired/in time series) | N/A |

| | |
|---|---|
| Additional sequence parameters (spectral width [Hz], number of spectral points, frequency offset), 2D FOV, matrix size, acceleration factors, sampling method | Spectral width: 5000Hz<br>Number of spectral points: 4096<br>Frequency offset: -2.3 ppm<br>Acceleration: None<br>Sampling method: Conventional |
| Water suppression method | VAPOR |
| Shimming method | FASTEST MAP; Siemens "Brain" B0 shim mode |
| Triggering or motion correction method | N/A |

### 3. Data analysis methods and outputs

| | |
|---|---|
| Analysis software | FID-A for data processing and LCModel for metabolite quantification |
| Processing steps deviating from quoted reference or product | N/A |
| Output measure | LCModel metabolite concentrations and ratios to Cr. |
| Quantification references and assumptions, fitting model assumptions | For LCModel, an internal unsuppressed water reference was used for metabolite quantification. The unsuppressed water scan was also used for eddy current correction. The LCModel default values for WCONC, ATTH2O, and ATTMET were used. |

### 4. Data quality

| | |
|---|---|
| Reported variables (SNR, linewidth (with reference peaks)) | SNR, LW |
| Data exclusion criteria | SNR<20; LW>10Hz |
| Quality measures of post processing model fitting (e.g., CRLB, goodness of fit, SD of residual) | SNR, LW, goodness of fit |

| Sample spectrum | Figure 1 |
|---|---|

**Supplementary Table S3**: Descriptive statistics for absolute metabolite concentrations and total creatine (tCr)-referenced ratios across healthy controls (HC), relapsing–remitting MS (RRMS), and progressive MS (PMS). Separate HC cohorts were used for RRMS and PMS comparisons to match age and sex distributions in each analysis.

| | RRMS vs HC | | | | PMS vs HC | | | |
|---|---|---|---|---|---|---|---|---|
| | PCC | | CSWM | | PCC | | CSWM | |
| | HC | RRMS | HC | RRMS | HC | PMS | HC | PMS |
| Absolute Metabolite Concentration | | | | | | | | |
| tNAA | 10.8(1.5) | 11.8(2.0) | 11.6(0.9) | 11.3(1.4) | 11.0(1.3) | 11.8(1.7) | 11.6(1.1) | 10.7(1.7) |
| mI | 6.0 (0.8) | 6.9(0.9) | 7.0(0.8) | 8.4(1.4) | 6.0(0.9) | 6.8(0.9) | 7.2(0.8 | 8.6(1.8) |
| tCho | 1.1(0.2) | 1.2(0.2) | 1.9(0.2) | 2.0(0.3) | 1.1(0.2) | 1.21(0.2) | 1.9(0.2) | 0.3(0.0) |
| Glx | 10.7(1.4) | 11.7(2.1) | 7.2(0.8) | 7.6(1.0) | 10.5(1.3) | 11.7(2.0) | 7.0(0.6) | 7.0(1.5) |
| tCr | 7.7(0.8) | 8.3(1.1) | 7.0(0.5) | 7.3(0.7) | 7.6(0.8) | 8.3(0.8) | 6.9(0.5) | 7.1(1.0) |
| GSH | 0.9(0.2) | 0.8(0.4) | 1.0(0.1) | 1.0(0.2) | 1.0(0.1) | 0.9(0.4) | 0.9(0.1) | 1.0(0.3) |
| mI/ tNAA | 0.7(0.1) | 0.7(0.1) | 0.8(0.1) | 1.0(0.3) | 0.6(0.1) | 0.7(0.1) | 0.8(0.1) | 1.0(0.2) |
| Ratio to tCr | | | | | | | | |
| tNAA/ tCr | 1.4(0.2) | 1.4(0.1) | 1.7(0.1) | 1.6(0.2) | 1.5(0.1) | 1.5(0.1) | 1.8(0.1) | 1.6(0.1) |
| mI/tCr | 0.8(0.1) | 0.9(0.1) | 1.0(0.1) | 1.2(0.2) | 0.8(0.1) | 0.8(0.1) | 1.1(0.1) | 1.3(0.2) |
| tCho/ tCr | 0.2(0.0) | 0.1(0.0) | 0.3(0.0) | 0.3(0.0) | 0.2(0.0) | 0.2(0.0) | 0.3(0.0) | 0.2(0.0) |
| Glx/ tCr | 1.4(0.1) | 1.4(0.2) | 1.0(0.1) | 1.0(0.1) | 1.4(0.1) | 1.4(0.1) | 1.0(0.1) | 1.0(0.1) |
| GSH/ tCr | 0.1(0.0) | 9.3E-02(0.0) | 0.1(0.0) | 1.41E-01(0.0) | 0.1(0.0) | 0.1(0.0) | 0.1(0.0) | 0.1(0.0) |
